# Parametric Bayesian Modelling of Tuberculosis Mortality determinants and Facility level heterogeneity effect using Gamma and Gaussian shared frailty techniques

**DOI:** 10.1101/2022.12.21.22283771

**Authors:** Isaac Fwemba, Veranyuy D. Ngah, Motlatsi Rangoanana, Llang Maama, Sele Maphalale, Mabatho Molete, Retselisitsoe Ratikoane, Modupe Ogunrombi, Olawande Daramola, Peter S. Nyasulu

## Abstract

**Background:** In a normal regression analysis for determinants of TB outcomes, assumptions that the sample is homogenous is made. This model does not account for the overall effect of unobserved or unmeasured covariates. This study aims to quantify the amount of heterogeneity that exists at community level, and to ascertain the determinants of TB mortality across all the catchment areas in Lesotho.

**Methods:** This was a retrospective record review of patients on TB treatment registered between January 2015 to December 2020 at 12 health care facilities in the district of Butha Buthe, Lesotho. Data collected from patient medical and statistical analysis was performed using R and INLA statistical software. Descriptive statistics were presented using frequency tables. Differences between binary outcomes were analysed using Person’s *X*^*2*^ test. Mixed effect model with five Bayesian regression models of varying distributions were used to assess heterogeneity at facility level. Kaplan-Meier curves were used to demonstrate time-to-death events

**Results:** The total number of patients included in the analysis were 1729 of which 70% were males. And half of them were employed (54.2%). Being over 60 years (HR: 0.02, Cl: 0.01-0.04) and having a community health worker as a treatment contact person (HR: 0.36, Cl: 0.19-0.71) decreased the risk of dying. Miners had 1.73 times increased risk of dying from TB (HR: 1.73, Cl: 1.07-2.78). The frailty variance was observed to be very minimal (<0.001), but significant indicating heterogeneity between catchment areas. Although similar hazard ratios and confidence intervals of covariates are seen between Gamma and Gaussian frailty log-logistic models, the credibility intervals for the Gamma model are consistently narrower.

**Conclusion:** The results from both Gamma and Gaussian demonstrate that heterogeneity affected significance of the determinants for TB mortality. The results showed community level to significantly affect the risk of dying indicating differences between catchment areas.

**Highlights:**

1. Reports of being employed as a miner associated with higher TB mortality is worrying. This finding may help authorities in Lesotho and the Southern African region to design health strategies that can target miners and those living within the mining catchment areas
2. The use of community health workers and close relatives reduced the risk of dying among TB patients. This is a key factor that can be considered in designing effective TB interventions in Lesotho. Ensuring that each patient is assigned a community health worker may reduce mortality.
3. The risk of death was significantly higher in treatment phase 2 among patients with pulmonary TB compared to patients in treatment phase 1 and among those with extra pulmonary TB

**Strength of the study:**

4. The study is based on mixed effect models with varying distribution for the frailty parameter. Selecting baseline hazard distribution is based on how the Lesotho data fits the model better and not based on historical practice that is not motivated by current data.
5. Presence of heterogeneity at a facility level means that interventions meant to improve TB treatment outcomes must be taken into consideration seriously.

## Background

Tuberculosis (TB) is a main leading causes of mortality worldwide from a single infectious disease (1). The highest burden of the disease is found in sub-Saharan Africa with 25% of worldwide incident cases in 2020 as reported by the World Health Organization (1,2).

TB control is a serious public health priority in many low and middle income countries (LMIC) as death rates among patients on TB treatment is still increasing globally (3,4). Several factors affecting survival of patients are missed in the TB treatment programs. Many studies assessing risk factors for survival of TB patients use the survival time statistical analysis which does not take into consideration the natural variability of the different environments among the patients (5).

In a normal regression model of tuberculosis treatment outcome in a specific population, the assumption that the sample is homogenous is made. Not only is this assumption unrealistic, but it is also impractical. This is because there are certain demographic differences among the patients which researchers are unaware of (6,7). In most studies, the burden of obtaining all the important explanatory variables causes a limitation due to important covariates being missed. This results in unwanted heterogeneity or clustering effect in the study (3,4).

Determining community level (frailty parameter) clustering effect is not new in biomedical research, but requires that the survival times for participants be independent and identically distributed (8). This assumption has been violated in practice, especially when data being analyzed involves participants that are connected on account of their relation within a family member (9), or by sharing community and environment factors (8,14). Similarly, other studies on TB outcomes, have shown that cluster levels factors (facility level) may have significant effects on mortality (10). Regrettably, community effects are normally not accounted for in studies assessing determinants of health outcomes such as TB. The use of frailty models are only being used recently in TB mortality studies (11,12).

Determinants of this nature represent the cumulative effect of unobserved or unmeasured covariates that may reflect impacts of environmental and socio-cultural factors. Various statistical methods are used to evaluate the size of the effect of these unobserved factors which have been known to act either multiplicatively or additively on the baseline hazards (8,13). Hougaard (14), and Klein (15), modelled the dependence of the covariate structure via frailty model of an assumed parametric distribution. Woya and Jabir et al., modelled the baseline hazard function using a Gamma shared frailty model and concluded that there was no heterogeneity for death in patients treated in different hospitals (11, 12). This study found that covariates such weight, age, extra pulmonary type of TB, and HIV status of TB patients were significant risk factors associated with death status among TB patients.

In practice modelling of frailty parameter decision has been done on the basis of traceability of the frailty function and availability of appropriate software (16) to implement these models and, not necessarily on their scientific merit. Lack of scientific evidence to support such decisions has affected appropriate inference needed to correctly estimate parameters needed to inform targeted TB interventions. This study aimed i) to examine how well a proposed distribution fits the data before any inference can be drawn, ii) to quantify the amount of heterogeneity existing at community level in Butha Buthe, Lesotho, which may bias mean estimates with corresponding standard deviations and consequently the credible intervals and lastly to iii) to identify determinants of TB mortality across all the catchment areas in Butha Buthe Lesotho.

## Methods

### Data Sources and Variables

This was a retrospective study conducted in ten primary care facilities and two secondary level hospitals in Butha-Buthe district. TB data were gathered from records of adult TB patients registered in health care facilities from 12 regions which includes (Butha-Buthe Government hospital (BBGH), Seboche Hospital, Boiketsiso, Linakeng, Makhunoane, Motete, Muela, Ngoajane, Rampai, St Paul, St Peters and Tsime clinics). A trained research coordinator supervised the process of data collection and ensured effective data quality controls. The current data contained parameters on 1729 TB patients. These patients were diagnosed with TB between the period of January 2015 and December 2020. Data collected included patients’ demographics (sex and age of the participant), TB outcomes, facility, TB treatment category, medication start date, medication completion date, phase of treatment, treatment contact group, and occupation.

To ensure data quality, we conducted a verification exercise by going through each facility’s records to ensure all erroneous entries were corrected or removed. We also verified TB outcomes (cure, death, lost to follow-up and transfer) and performed data consolidation. The outcome variable was defined as survival time in days of patients who were diagnosed with TB. Those who died because of TB were patients classified as having had the event and assigned to the number ‘1’. Patients who either dropped out of the study or transferred out were censored using mechanism as illustrated in equation (8) and were assigned the number ‘0’.

### Statistical Analysis

#### Outcome variable

Outcome variable was defined death from TB measured as survival time in days after TB diagnosis. Patients deemed to have had the event were assigned the number ‘1’.

#### Analytical approach

Several parametric regression models were fitted using the Bayesian frameworks. Fifteen models were specified for this work. The first five were Bayesian regression models (loglogistic, lognormal, exponential, Weibull and Gompertz) specified and fitted with the assumption that community heterogeneity (frailty) is homogenous across the catchment areas.

We used the proportional hazards model as shown

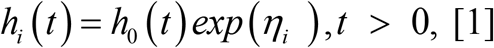

where the baseline hazard is represented by *h*_0_ (·), and the predictors by *η*_*i*_,

In this analysis, assumptions that the data follow the right censoring mechanism were made, where patients who died from TB are said to have experienced the event while all others (lost to follow-up, dropouts, and transfer outs) were censored (have not had the event of interest).

#### Exponential Distribution

We assumed that the hazard function is constant over time. The hazard of death at any time after the time origin of the study is then the same irrespective of the time change. This property of the exponential distribution is called “loss of memory” which requires that the age of the person does not affect future survival. Let it be the survival time that follows exponential distribution with parameter λ. Therefore, the probability density function (pdf) of t is

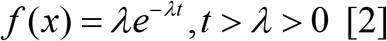

Hence *h*(*t*) = *λ*

#### Gompertz Distribution

The probability density function of the Gompertz distribution is shown below

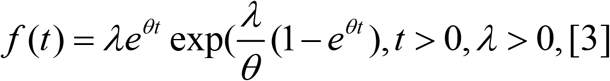

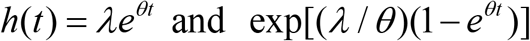

#### Weibull Distribution

The Weibull distribution is suitable for modeling data with monotone hazard rates that increase or decrease exponentially over time. For Weibull regression, λ is the scale parameter and γ is a shape parameter, and its probability distribution function shown below

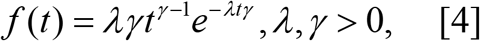

Expressing the latent field x, in terms of the predictor *η*_*i*_, the standard Weibull regression is expressed *h* = *λ*_*t*_*t*^*λ*− 1^*exp*(*η*)

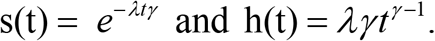

#### The Log-Logistic Distribution

The Weibull hazard is a monotonic function of time which is a limitation. However, situations can arise in which the hazard function changes direction. In this situation, log-logistic is a preferable model.

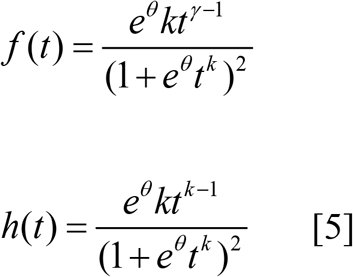

For 0 ≤ *t* < ∝ and 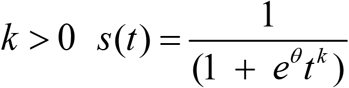

#### The Log-Normal Distribution

The log-normal distribution can be used as a model for survival data since it can be defined for random variables. A random variable T is said to have a log-normal distribution with parameters *μ* and *δ* log T, with a normal distribution *μ* and variance *δ*. Hence, the probability density function of T is as follows

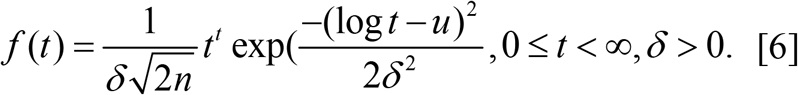

The survivor function of the log-normal distribution is

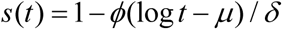

Where *ϕ*(.) is the standard normal distribution function given by

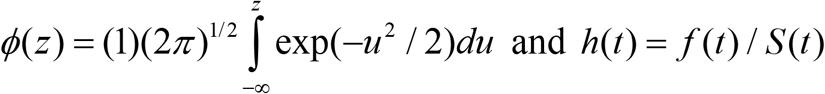

#### Prior Information

We specify a Gaussian distribution with hyper parameter as priors for all regression coefficients and the Gamma distribution *θ*= (*θ*_1_, *θ*_2_) as priors for frailty parameter. Similarly, hyper parameters were assigned a Gamma distribution Γ(*a, b*) with 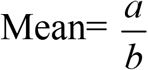 and 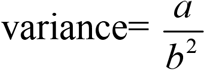 as priors for *λ*. Flat priors were also assigned *τ*∼ Γ(1, 1)

#### Gamma and Gaussian Frailty Models as conditional survival models

We modelled and analyzed the time to death of TB patients, by considering several parametric and frailty survival models. Frailties are random effects accounting for association or unobserved heterogeneity of clustered survival data. In a shared frailty model, the lifetimes of a group of observations in the same cluster share the same level of frailty (8,12,17,18). An advantage of the parametric models is that they are better fit models over Cox when the shape of the hazard is known. Thus, we fitted 10 Bayesian regression models in addition to the first five fitted. These models included a Gamma and Gaussian shared frailty term with the assumption of a significant unobserved effect (presence of heterogeneity). Analysis was conducted on each model through the Bayesian approach for all the data sets.

#### Shared Frailty Model

We assume that there are m clusters and each cluster *i* has *n*_*i*_ observations with a total sample size 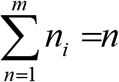. Let 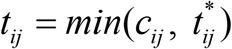 be the observed failure time with a right censoring scheme for the *j*^*th*^ (*j* = 1, 2, …, *n*_*i*_) observation in the *i*^*th*^ cluster, and *c*_*ij*_ be the censoring time; it is assumed that the failure time 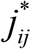 and censoring time *c*_*ij*_ are independently distributed random variables(17,19). In addition, let the random variable *v*_*i*_ be the frailty effect of the *i*^*th*^ cluster with known distribution function *f*_*v*_ (*v*_*i*_). Conditional on the frailty variable *v*_*i*_ and covariates *x*_*ij*_, the survival function *s*_*ij*_ (. | *x*_*ij*_ *v*_*ij*_) at time *t* of the *j*^*th*^ observation of cluster *i*, is given by (18, 20, 21);

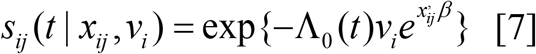

where, Λ_0_ (*t*) is the cumulative baseline hazard function which needs to be specified for generating survival data, and *β* represents the unknown regression parameters. For invertible Λ_0_ (*t*), the failure times can be computed by

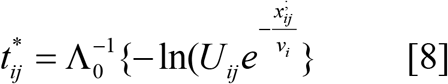

where, *U*_*ij*_ ∼ *U* (0, 1) and 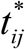 is the failure time of member *j* of cluster *i*. Then the observed censoring indicator *δ*_*ij*_ is equal to 1 if 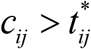, and 0 otherwise. Conditional on the frailty term *ν*_*i*_ and the covariate *x*_*ij*_, the shared frailty model at time *t* of the *j*^*th*^ observation of cluster *i* is defined in terms of a conditional hazard model

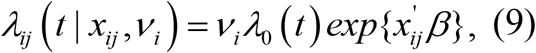

where *λ*_0_ (.) is the baseline hazard function, and β is a vector of the regression parameters. If *n*_*i*_ = 1 for *i* = 1, 2, …, *m*, then the survival function in Expression (1) and the hazard function in Expression (2) reduce to a proportional hazard model (18). The frailty effects *ν*_*i*_, *I* = 1, …, *m* are assumed to be independent and identically distributed random variables with distribution functions such as log-normal, positive stable distribution, Gamma and inverse Gaussian distributions (19, 20).

Due to their computational convenience, the Gamma and the inverse Gaussian distributions were used as the frailty distributions for this study

#### A Shared Gamma Frailty Model

The two parameter Gamma density function for the frailty term *v*_*i*_ with shape parameters k and scale parameter λ are given by

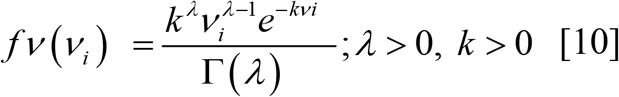

The Laplace transformed version of this density has a form (8):

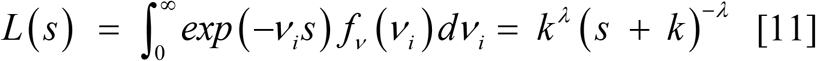

The solutions of the first and second partial derivatives of the Laplace function L(.) with respect to s equal to 0 give the mean and variance of the frailty term 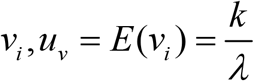 and 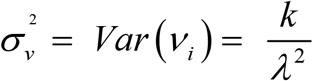, respectively. For identifiability reasons, we set the mean *μ*_*ν*_ of the frailty term equal to 1 and the variance of the frailty term 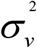 becomes 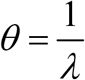, which is similar to the one-parameter Gamma distribution. The shared Gamma frailty model (conditional hazard) for individual *j* in cluster *i* is then given by:

For identification, we set the mean μν of the frailty term equal to 1 and the variance of the frailty term 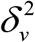 becomes 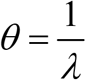, which is similar to the one-parameter Gamma distribution. The shared Gamma frailty model (conditional hazard) for individual in cluster *i* is then given by:

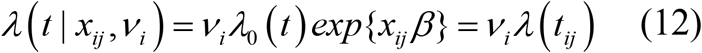

#### The Inverse Gaussian Frailty Model

The inverse Gaussian (inverse normal) distribution was introduced as a frailty distribution alternative to the Gamma distribution. Like the Gamma frailty model, simple closed-form expressions exist for the unconditional survival and hazard functions, which makes the model more plausible. The probability density function of an inverse Gaussian distributed frailty random variable *ν*_*i*_ with parameters μ > 0 and *α* > 0 is given by (18-22),

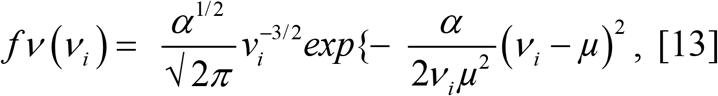

where, the Laplace transform of this function has a form

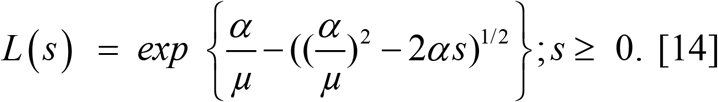

#### Reporting estimates from statistical analysis presented

Frequencies and associated proportions were determined for categorical measures and compared using the Chi-square test. The Kaplan–Meier test was used to determine differences in survival by sex, age category and catchment areas.

Factors associated with mortality were assessed using the integrated nested Laplace approximation (INLA), where univariate and multivariate shared Gamma and Gaussian frailty models were fitted. Estimates generated from INLA were exponentiated to obtain posterior summaries.

Bayesian summaries were reported as hazard ratios with their associated 95% confidence intervals. INLA accounted for unmeasured variables and allowed for the comparison of frailty variances between patients, region, and tumor extent. A significant frailty variance with a value <1 suggests a higher rate for the event (or shorter survival times) than would be predicted under the basic Weibull model, while a value >1 suggests a lower rate denoting a longer survival time. A 95% confidence interval excluding one was determined as significant. Model fit was assessed using the deviance information criterion (DIC) and Watanabe Information. All statistical analyses were performed using R for frequentist approach, while Bayesian Approach was implemented using deterministic based approach known as INLA, a variant of R open-source software. The assumption of a two-sided test at 5% significance level was used to evaluate for significance of the results. Before running the models, we scaled the survival time to the maximum of 1. We standardized time to avoid convergence challenges

## Results

A total of 1792 records were reviewed and analyzed. After censoring of patients whose outcome data were unavailable, 1729 patient records were included in the analysis. In Table 1 below, we provide a summary of the covariates included in the analysis. There are evident disparities by catchment areas (facility names), with Seboche, and BBGH catchment areas having the highest number of TB patients accounting for 50%, and 22.9%, respectively. Phase 1 of treatment (64.6%), having pulmonary TB (77.1%) were the most common observed characteristics in these areas. In general, there were 19.1% TB related deaths and 80.9% TB patients who survived the episode, and 70.1% of the participants were males.

**Table 1:** Distribution of characteristics of the study participants.

| **Variables** | **Frequency (%)** |
| --- | --- |
| **Sex** |  |
| Female | 513 (29.7%) |
| Male | 1213 (70.3%) |
| **Employment status** |  |
| Employed | 935 (54.2%) |
| Unemployed | 791 (45.8%) |
| **Drug Resistance** |  |
| Drug Res | 213 (12.3%) |
| Drug Susceptible | 1513 (87.7%) |
| **Age category** |  |
| <20 Yrs. | 69 (4.00%) |
| ≥60 Yrs. | 378 (21.9%) |
| 20-59 Yrs. | 1279 (74.1%) |
| **Occupation Category** |  |
| Employed non-mine worker | 390 (22.6%) |
| Mine Worker | 549 (31.8%) |
| Unemployed | 787 (45.6%) |
| **TB Category** |  |
| Extra pulmonary TB | 403 (23.3%) |
| Pulmonary TB | 1323 (76.7%) |
| **Treatment contact group** |  |
| CHW | 267 (15.5%) |
| Fam Mem | 866 (50.2%) |
| Friend | 152 (8.81%) |
| Unknown | 441 (25.6%) |
| **Phase Of treatment** |  |
| Phase 1 | 1103 (63.9%) |
| Phase 2 | 623 (36.1%) |
| **Facilities** |  |
| BBGH | 380(22.02%) |
| Boiketsiso | 77(4.46%) |
| Linakeng | 45(2.61%) |
| Makhunoane | 33(1.91%) |
| Motete | 8(0.46%) |
| Muela_Botha | 24(1.39%) |
| Ngoajane | 47(2.72%) |
| Rampai | 24(1.39%) |
| Seboche | 884(51.22%) |
| St_Paul | 89(5.16%) |
| St_Peters | 57(3.3%) |
| Tsima | 58(3.36%) |

### Comparison of Survival Curves

The survival distributions of time to death of the TB patients were estimated for each group using the Kaplan–Meier (KM) method to compare the survival curves of two or more groups. The Kaplan–Meier estimated survival curves in Figure 1-10, highlight the overall estimated survival function using distinct groups of covariates. The overall estimated survival curve for the time to death of TB patients is shown in Figure 1.0. In addition, the survival curves of TB patients were different for the different phase of treatment, treatment contact, drug resistance, category of TB, age category and catchment areas (facility) (Figures 1-5, and Figures 8-10) whereas amongst gender, and treatment category there were no clear differences.

**Figure 1:**
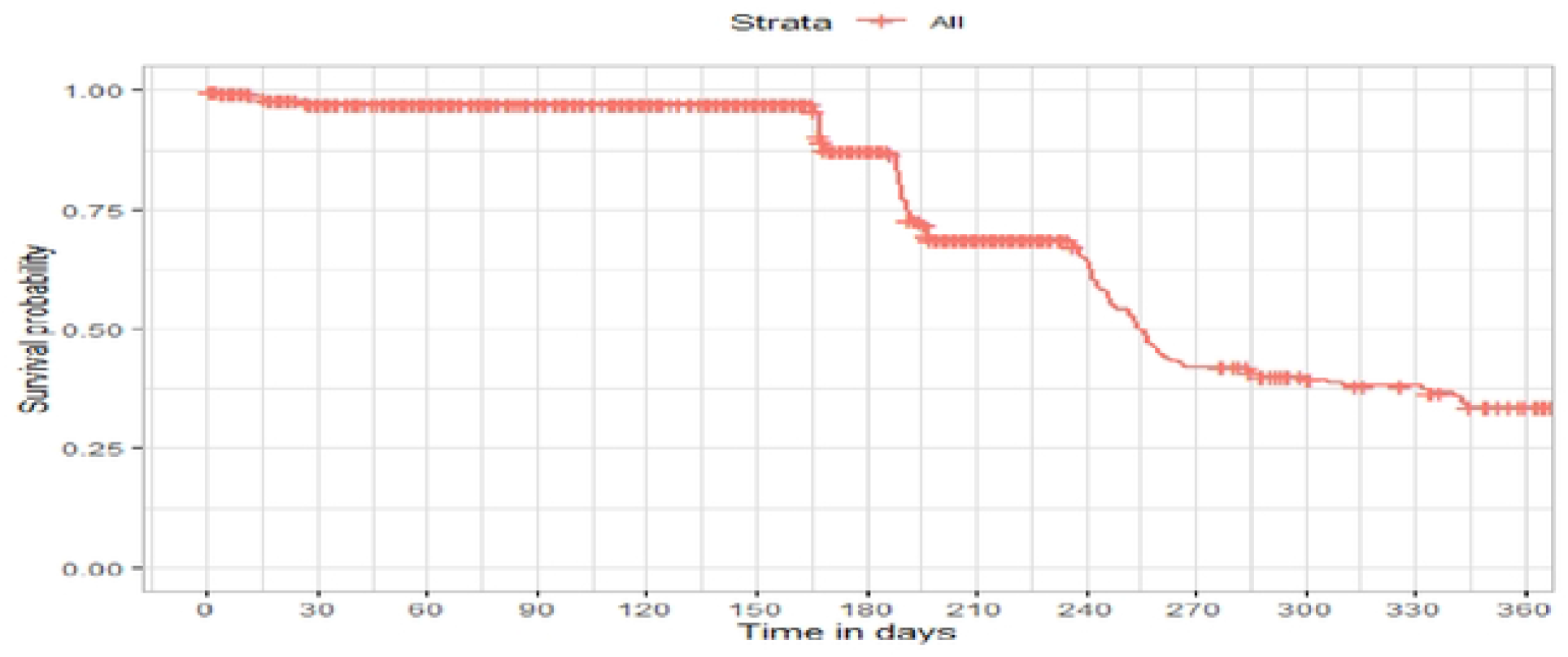
Overall Survival

**Figure 2:**
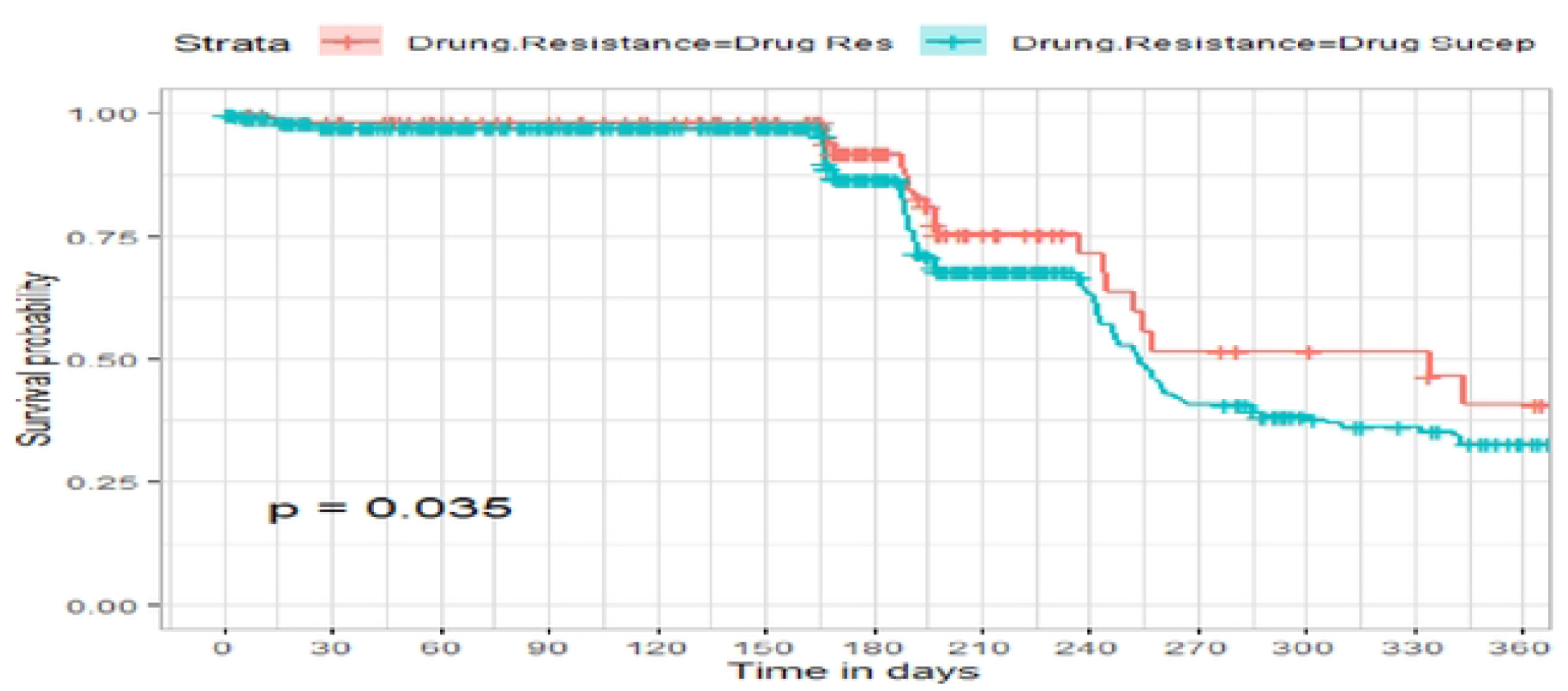
Survival curves for Drug Resistance

**Figure 3:**
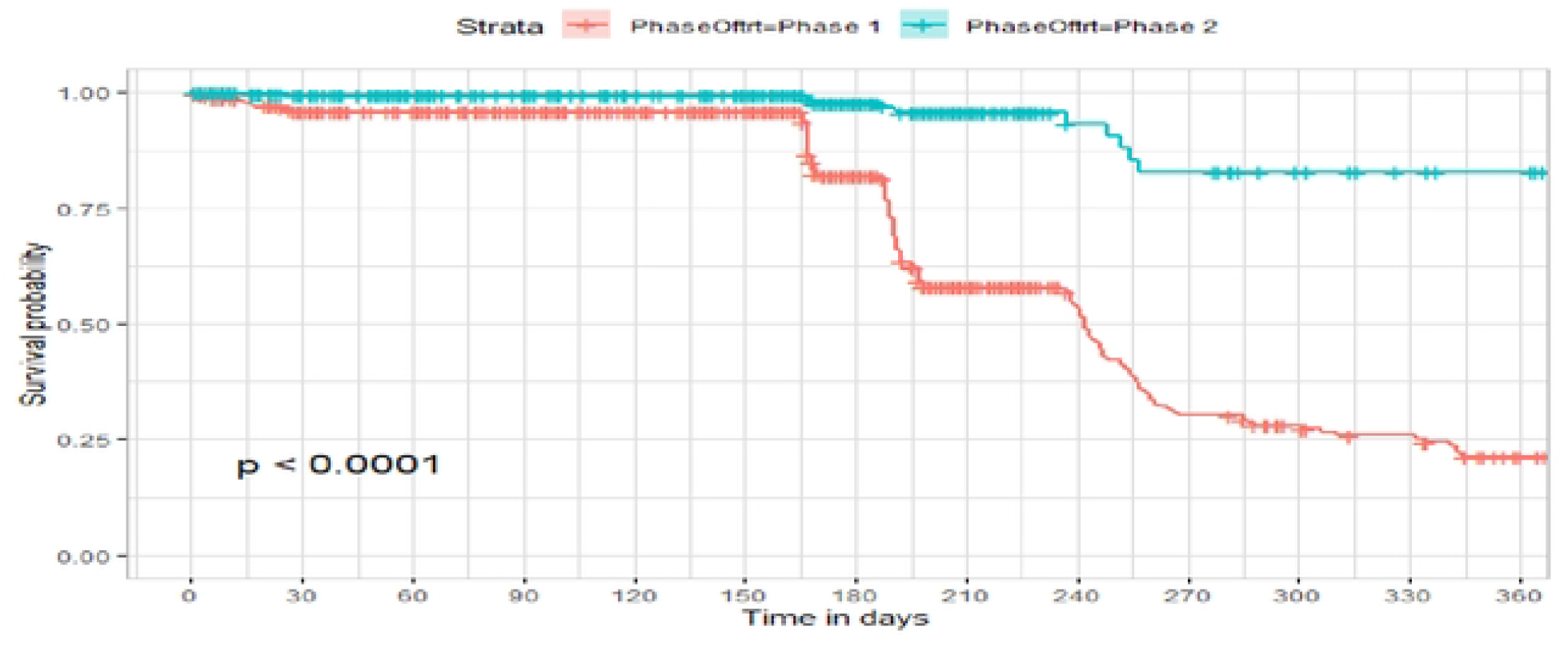
Survival Curves for Phase of treatment

**Figure 4:**
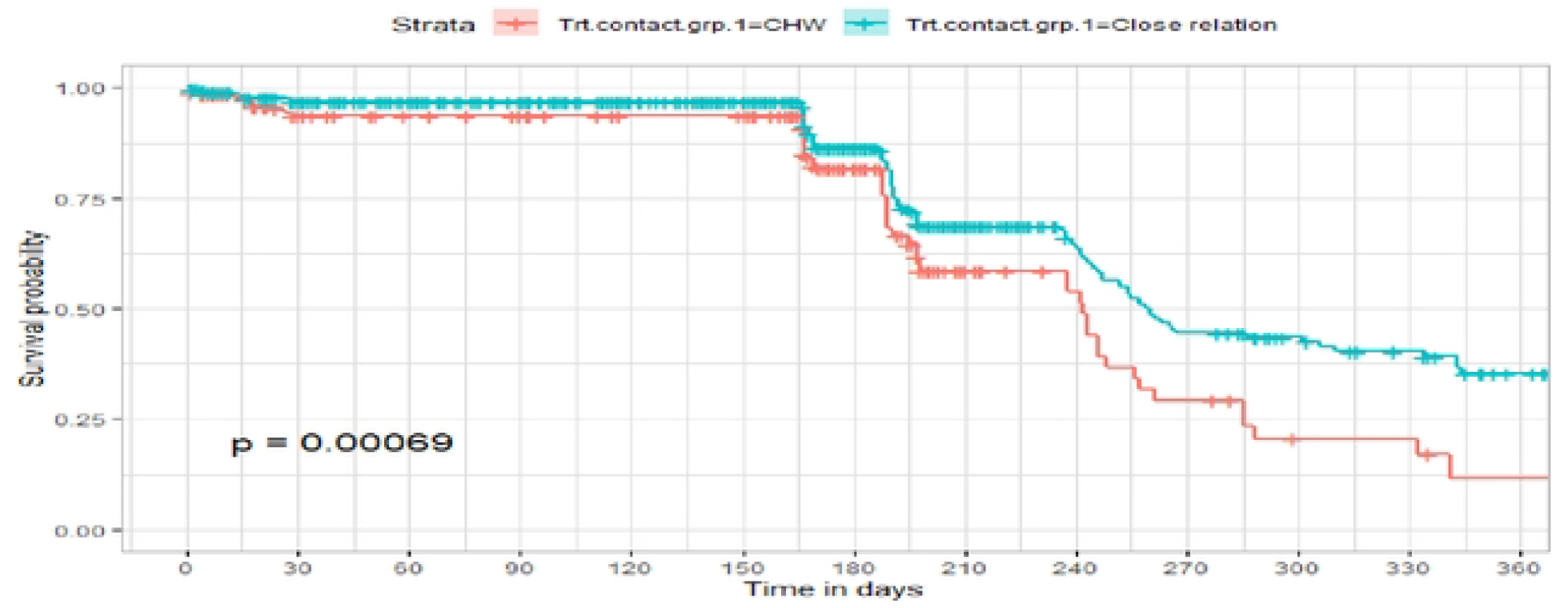
Survival Curves for treatment contact

**Figure 5:**
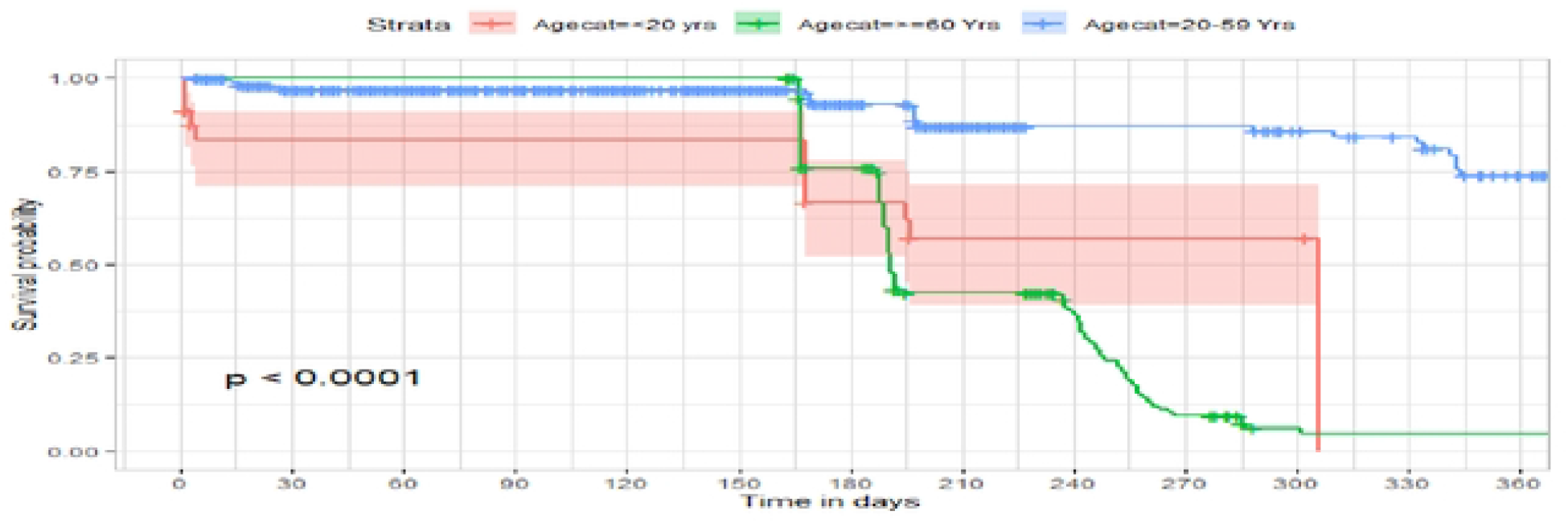
Survival curves for Age category

**Figure 6:**
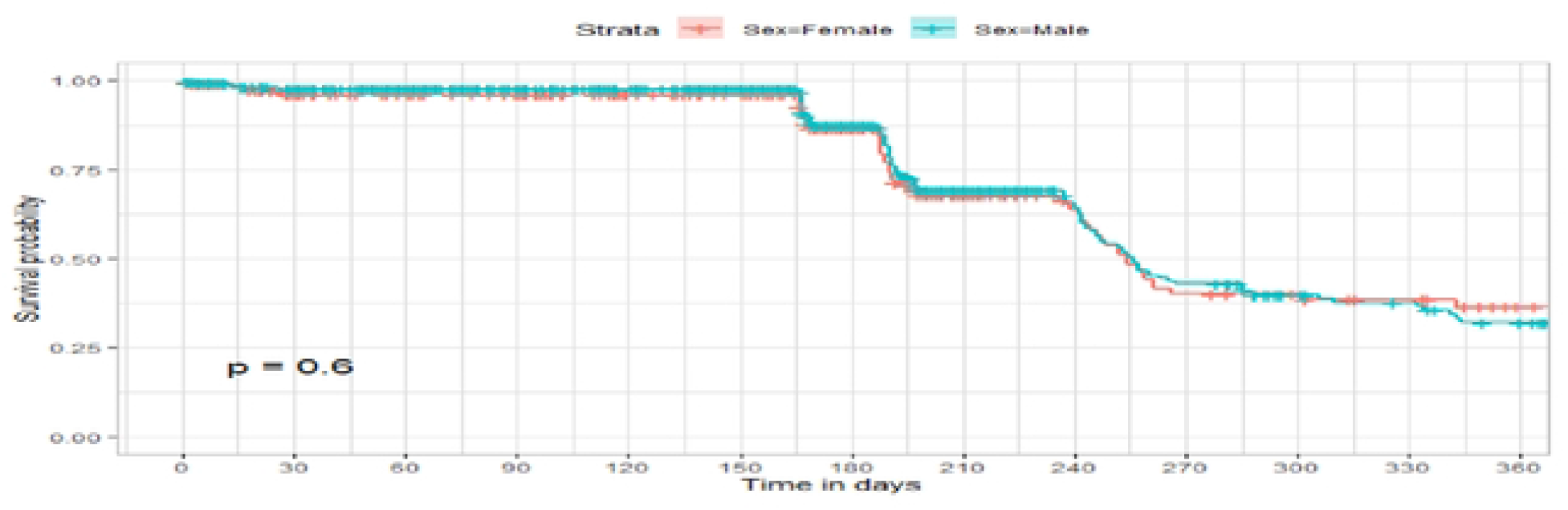
Survival Curves for Sex

**Figure 7:**
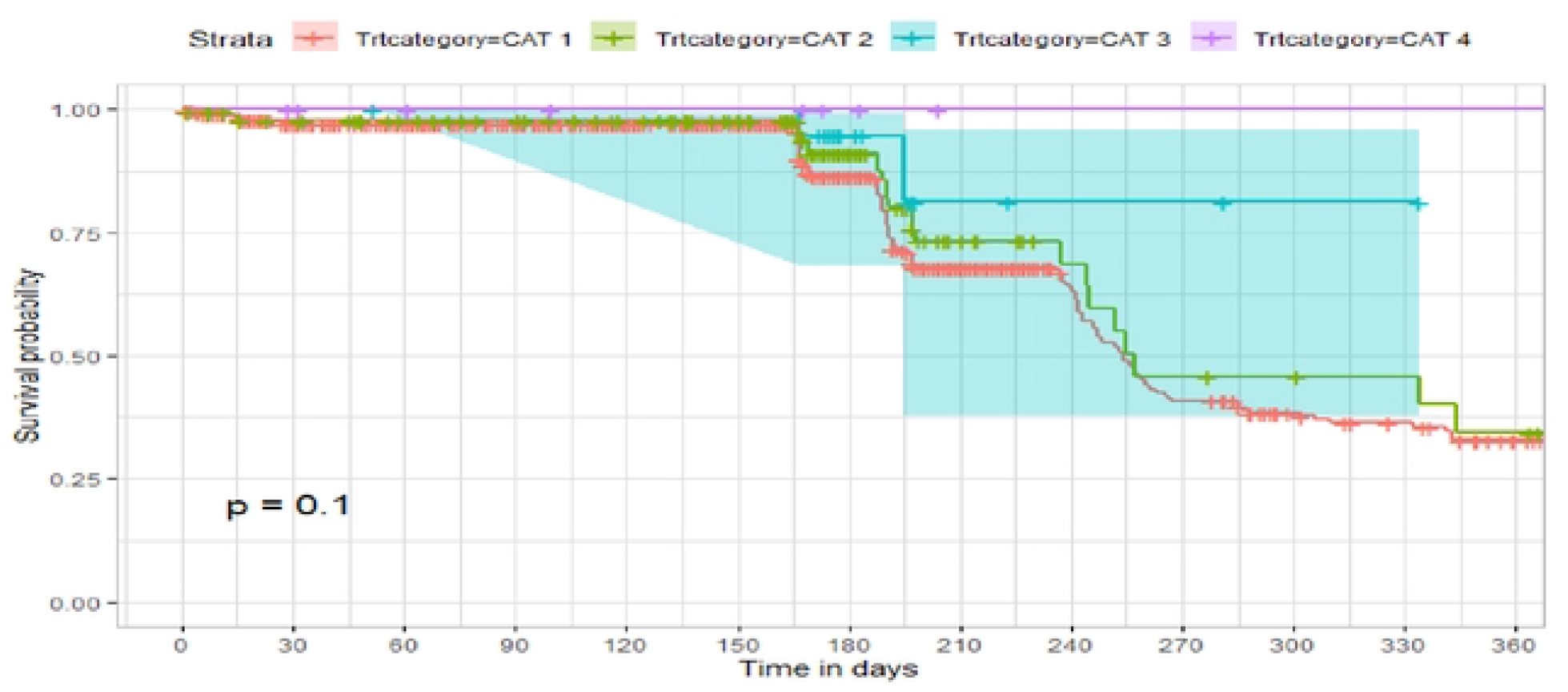
Survival Curves for treatment category

**Figure 8:**
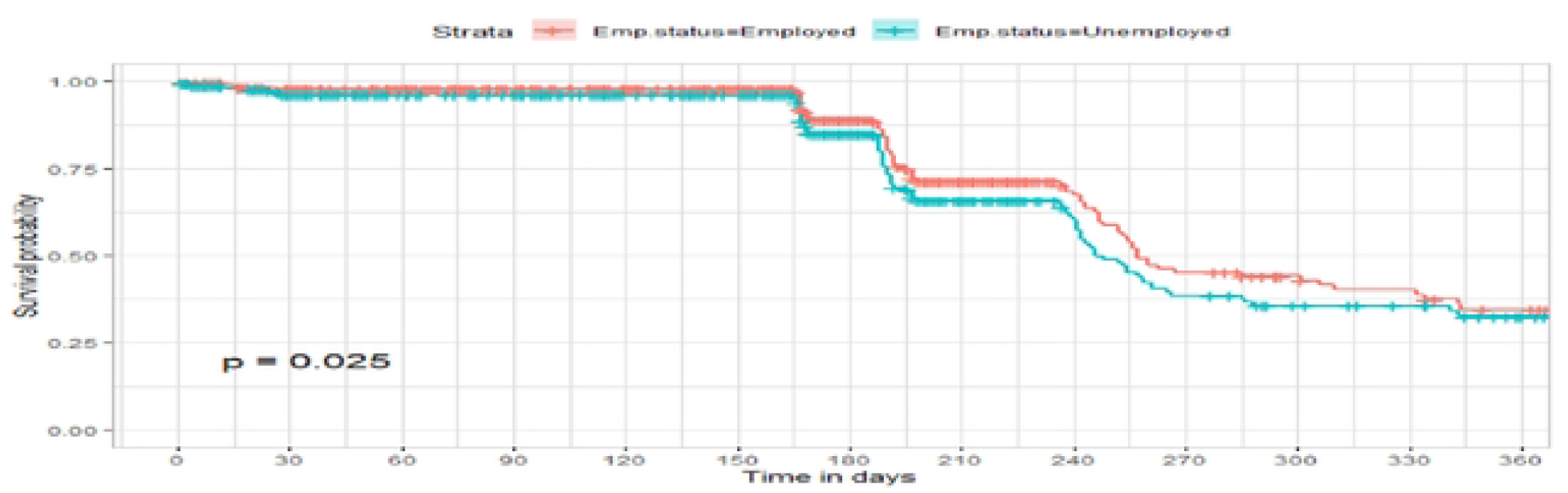
Survival Curves for Employment Status

**Figure 9:**
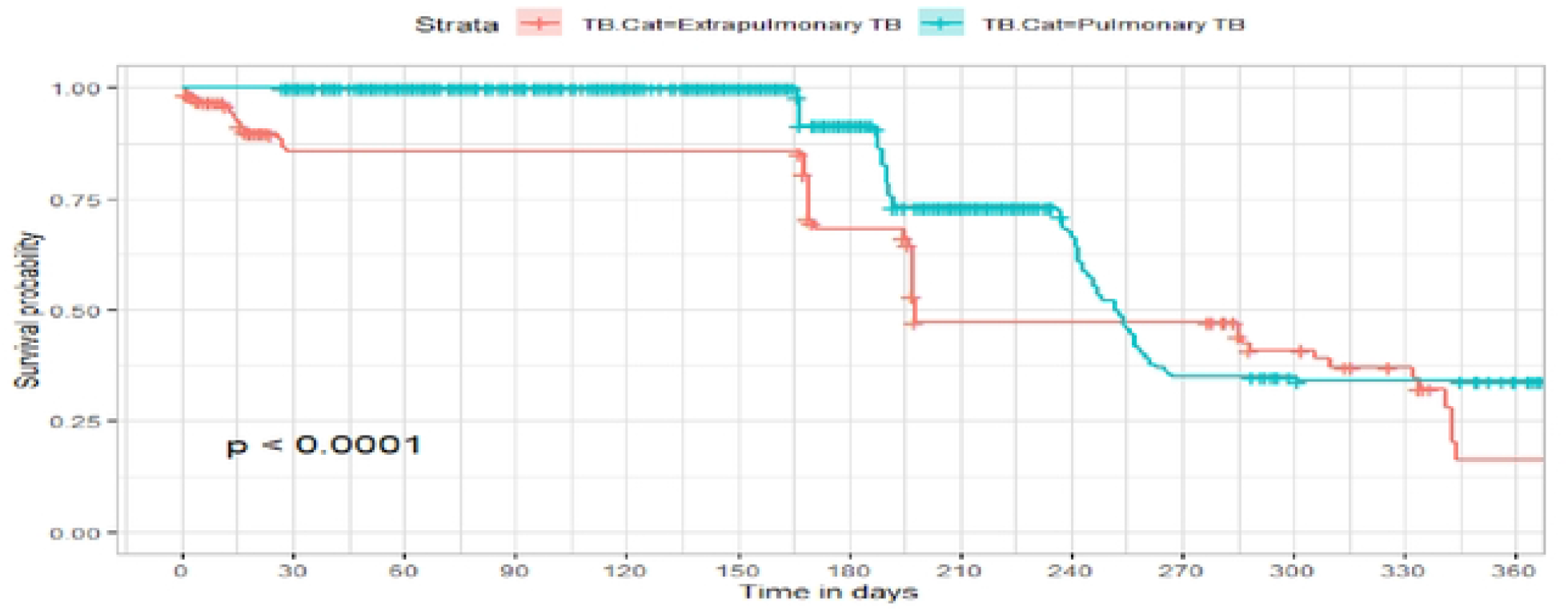
Survival Curves for TB. Category

**Figure 10:**
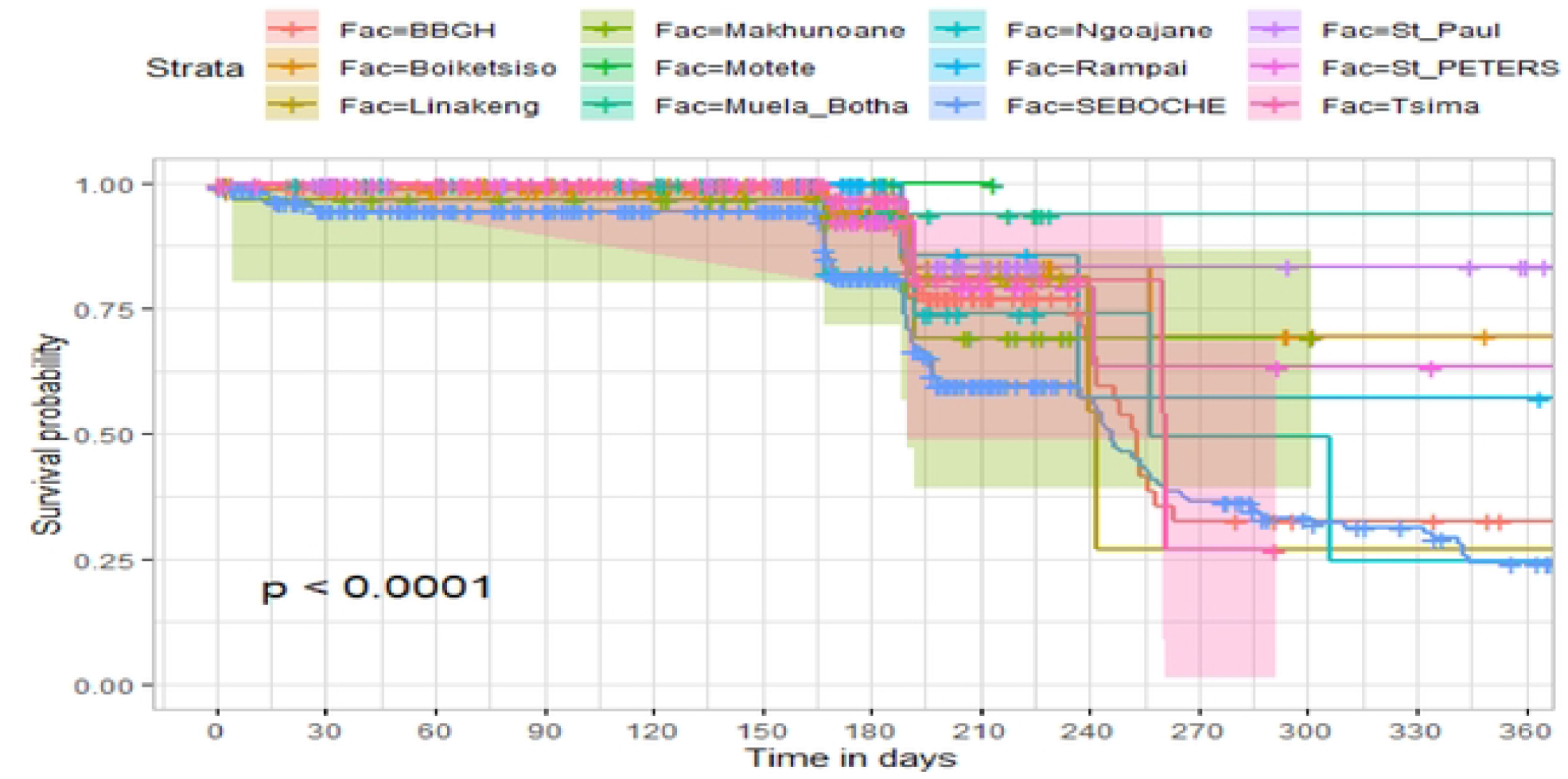
Survival Curves for Facility

The observed differences in survival experiences in different patient groups was also assessed using the log-rank test. There was a significant survival time difference at the 5% significance level among phase of treatment, treatment contact, drug resistance, category of TB, age category and facility for TB patients (Table 2). The results complement the graphical presentation shown in the figures above.

**Table 2:** Chi-square estimate (*x*^2^) and p-value of log-rank tests for comparing the survival experience of TB patients from randomly selected hospitals in Lesotho.

| No. | Covariate | $\chi^2$ | p |
| --- | --- | --- | --- |
| 1 | Sex | 0.26 | 0.61 |
| 2 | Treatment category | 6.44 | 0.095 |
| 3 | Age category | 281.92 | <0.001 |
| 4 | Treatment contact group | 24.62 | <0.001 |
| 5 | Employment status | 5.04 | 0.025 |
| 6 | Phase of treatment | 125.34 | <0.001 |
| 7 | Drug resistance | 4.47 | 0.035 |
| 9 | TB category | 37.25 | 0.001 |
| 10 | Facility | 57.48 | <0.001 |

### Model Assessment and Evaluation

To identify sets of covariates that have the potential influence to be included in the linear components of a multivariable model, literature review from similar studies was conducted (9-12). Given, fewer covariates that were abstracted from the database, we included all in the multivariable analysis. We performed multivariable survival analysis by assuming exponential, Weibull, lognormal, and Log logistic distributions for baseline hazard functions and varied the two frailty distributions. Covariates with unambiguous estimates such as category of treatment and drug resistance were dropped from all the models.

Exponential, Weibull, Gompertz, lognormal and log logistic regression models without frailty were fitted first using INLA framework. We varied the distributions of the frailty term to determine the best distribution for the frailty parameter being estimated using TB data from Butha Buthe district, Lesotho. The Gamma and inverse-Gaussian distributions are the most common and widely used distributions cited in the literature for modeling the frailty effect (8,16). Thus, we used the data to determine the appropriate frailty distribution, as shown in (Table 3) below.

**Table 3:** Model Assessment and Evaluation.

| No | Distribution | WAIC Values |
| --- | --- | --- |
| **a. Fixed Effect** |  |  |
| 1 | Weibull | -13.49 |
| 2 | **log logistic** | **-126.50** |
| 3 | Exponential | 40.95 |
| 4 | lognormal | -121.63 |
| 5 | Gompertz | 43.19 |
| **b. Gaussian distribution for frailty term** |  |  |
| 6 | Weibull | -69.40 |
| 7 | **Log logistic** | **-185.70** |
| 8 | Exponential | -29.92 |
| 9 | Lognormal | -175.18 |
| 10 | Gompertz | -28.07 |
| **c. Gamma distribution for the frailty term** |  |  |
| 11 | Weibull | -17428.47 |
| 12 | Exponential | 1702436.92 |
| 13 | **log logistic** | **-25587.79** |
| 14 | lognormal | -25576.55 |
| 15 | Gompertz | -25456.42 |

The model distribution and associated watanable akaike information criterion which is the generalization of the AIC for Gamma and Inverse-Gaussian shared frailty models are displayed in Table 3. Observing from the table above, model exponential is the worst performing model compared to all the other models with Gamma and Gaussian shared frailty terms. This is because it has the highest WAIC value. Log logistic, is the best performing model with or without the frailty term for both Gamma and Gaussian distribution. We can therefore conclude that the strongest and decisive model is the log logistic model with a Gamma shared frailty. The final analysis and results interpretation were based on the logistic regression model with a Gamma frailty term as shown in Table 4 below.

**Table 4:** Fixed effect models with Gamma frailty distribution.

|  | Weibull |  |  | Log logistic |  |  | Exponential |  |  | lognormal |  |  | Gompertz |  |  |
| --- | --- | --- | --- | --- | --- | --- | --- | --- | --- | --- | --- | --- | --- | --- | --- |
|  | HR | 2.50% | 97.50% | HR | 2.50% | 97.50% | HR | 2.50% | 97.50% | HR | 2.50% | 97.50% | HR | 2.50% | 97.50% |
| **Gender** |  |  |  |  |  |  |  |  |  |  |  |  |  |  |  |
| Female |  |  |  |  |  |  |  |  |  |  |  |  |  |  |  |
| Males | 0.91 | 0.69 | 1.22 | 0.89 | 0.6 | 1.33 | 0.91 | 0.68 | 1.22 | 0.92 | 0.62 | 1.37 | 1.30 | 0.99 | 1.72 |
| **Age Category** |  |  |  |  |  |  |  |  |  |  |  |  |  |  |  |
| <20Yrs |  |  |  |  |  |  |  |  |  |  |  |  |  |  |  |
| 20-59 yrs. | 0.87 | 0.54 | 1.45 | 0.85 | 0.43 | 1.63 | 0.92 | 0.57 | 1.53 | 0.98 | 0.52 | 1.84 | 0.87 | 0.53 | 1.43 |
| >=60 yrs. | 11.18 | 6.89 | 18.83* | 0.01 | 0.01 | 0.03* | 12.62 | 7.79 | 21.22 | 0.02 | 0.01 | 0.04 | 11.59 | 7.09 | 18.95 |
| **Treatment Contact** |  |  |  |  |  |  |  |  |  |  |  |  |  |  |  |
| HCW |  |  |  |  |  |  |  |  |  |  |  |  |  |  |  |
| CHW | 5.39 | 3.74 | 7.80* | 0.105 | 0.062 | 0.176* | 6.58 | 4.58 | 9.48* | 0.107 | 0.06 | 0.19* | 6.61 | 4.60 | 9.50* |
| Close relation | 1.94 | 1.43 | 2.67* | 0.355 | 0.235 | 0.532* | 2.03 | 1.49 | 2.79* | 0.367 | 0.24 | 0.57* | 2.07 | 1.51 | 2.83* |
| **Occupation** |  |  |  |  |  |  |  |  |  |  |  |  |  |  |  |
| Employed |  |  |  |  |  |  |  |  |  |  |  |  |  |  |  |
| Mine Worker | 0.80 | 0.57 | 1.13 | 1.73 | 1.07 | 2.78* | 0.77 | 0.55 | 1.08 | 1.53 | 0.95 | 2.47 | 0.81 | 0.58 | 1.14 |
| Unemployed | 0.65 | 0.4 | 1.06 | 1.47 | 0.74 | 2.89 | 0.62 | 0.38 | 1.01 | 1.51 | 0.77 | 2.97 | 0.97 | 0.64 | 1.49 |
| **Employment status** |  |  |  |  |  |  |  |  |  |  |  |  |  |  |  |
| Employed |  |  |  |  |  |  |  |  |  |  |  |  |  |  |  |
| Unemployed | 1.5 | 0.93 | 2.47 | 0.64 | 0.32 | 1.27 | 1.55 | 0.96 | 2.54 | 0.59 | 0.3 | 1.16 | 1.49 | 0.95 | 2.34 |
| **Phase of Treatment** |  |  |  |  |  |  |  |  |  |  |  |  |  |  |  |
| Phase 1 |  |  |  |  |  |  |  |  |  |  |  |  |  |  |  |
| Phase 2 | 0.04 | 0.03 | 0.07 | 65.51 | 35.23 | 129.42* | 0.04 | 0.02 | 0.06* | 39.54 | 23.87 | 68.19 | 0.05 | 0.03 | 0.08 |
| **TB Category** |  |  |  |  |  |  |  |  |  |  |  |  |  |  |  |
| Extra Pulmonary |  |  |  |  |  |  |  |  |  |  |  |  |  |  |  |
| Pulmonary TB | 0.24 | 0.18 | 0.31 | 19.93 | 12 | 34.08* | 0.24 | 0.18 | 0.32* | 17.21 | 10.59 | 28.94 | 0.25 | 0.19 | 0.33 |

We estimated five models that assume that TB patients from the same region have uncorrelated survival risk. Furthermore, ten additional models were constructed to control for frailty effect at the regional level. Inclusion of the frailty term strategy enabled us to examine the effects of unobserved factor estimates of the known determinants for TB. Fixed effects results from the standard Weibull, Exponential, Lognormal, Gompertz and Log logistic models constructed to analyze the TB data are presented in Table 4

### Determinants of TB mortality in Lesotho

Table 6 shows the hazard rates of TB mortality in twelve health facilities considered in the study with a Bayesian log logistic regression modeling approach via a Gamma shared frailty. and presents estimates of fixed risk factors resulting from the models with the best fit. It is interesting to note that a male TB patient had risk of dying relative to female patient, however this was not statistically significant (HR=0.76 CI 0.52-1.12).

While being over 60 years, have been contacted by community health worker and close relatives reduced the hazard of dying. Being over 60 years (HR: 0.02, Cl: 0.01-0.04) and being contacted by community health workers (HR:0.36, Cl 0.19-0.71) were associated with lower hazard of dying. Those who contacted close family relatives were not significantly different from those who did not (HR:0.93, Cl:0.54-1.61), even though the effect was found to be reducing.

Interestingly, the effect of occupation on time of death was found to be linear. Those who were employed as miners were 1.73 times more at risk of dying from TB (HR:1.73, Cl 1.07-2.78). The effect was also positive for those who were unemployed but living in the same mining area (HR; 1.47, Cl: 0.74-2.89). This effect was however not statistically significant.

Analyzing the effect of employment status alone on mortality without stratifying for mining (miner and non-miner) masked the effect of occupation on death. The results showed a non-significant relationship (HR: 1.25, Cl: 0.64-2.45). The overall risk of death also reduced from 1.70 to 1.25. This finding show that being in employment or not, did not significantly affect TB mortality for those living in the study area, however working as a miner increased the risk of dying as reported above.

Surprisingly, unambiguous results were also noted. For instance, the effect of treatment category phase 2 (HR: 71.16; Cl: 37.4-142.96) increased the risk of dying relative to those in phase 1. This relationship was significant at p<0.05. Similarly, the effect of pulmonary TB (HR: 16.76, Cl: 10-28.81) on mortality was 19.93 times higher relative to those who were diagnosed with extra pulmonary TB

### Modeling heterogeneity using frailty Models. Explaining the Gamma parameter and the unobserved effects

Tables 5 and 6 provide results from the two survival mixed effect models fitted to control for unobserved effects (catchment region). Table 5 presents results for estimates from models with Gaussian distribution for the random shared parameter, while estimates in table 6 are those from models that included the Gamma parameter as the distribution of choice for the random or frailty term.

**Table 5:** Random effect with Gaussian distribution.

|  | Weibull frailty |  |  | log logistic |  |  | Exponential frailty |  |  | Lognormal frailty |  |  | Gompertz frailty |  |  |
| --- | --- | --- | --- | --- | --- | --- | --- | --- | --- | --- | --- | --- | --- | --- | --- |
|  | HR | 2.50% | 97.50% | HR | 2.50% | 97.50% | HR | 2.50% | 97.50% | HR | 2.50% | 97.50% | HR | 2.50% | 97.50% |
| **Gender** |  |  |  |  |  |  |  |  |  |  |  |  |  |  |  |
| Female |  |  |  |  |  |  |  |  |  |  |  |  |  |  |  |
| Male | 0.93 | 0.7 | 1.24 | 0.89 | 0.59 | 1.32 | 0.93 | 0.70 | 1.24 | 0.91 | 0.62 | 1.35 | 1.151 | 0.876 | 1.512 |
| **Age category** |  |  |  |  |  |  |  |  |  |  |  |  |  |  |  |
| <20Yrs |  |  |  |  |  |  |  |  |  |  |  |  |  |  |  |
| 20-59 Yrs. | 0.81 | 0.5 | 1.35 | 1.02 | 0.51 | 2.00 | 0.85 | 0.53 | 1.42 | 1.18 | 0.62 | 2.24 | 0.751 | 0.457 | 1.234 |
| >=60 Yrs. | 9.51 | 5.81 | 16.15 | 0.02 | 0.01 | 0.04* | 10.47 | 6.4 | 17.73 | 0.02 | 0.01 | 0.05 | 8.58 | 5.20 | 14.14 |
| **Treatment Contact** |  |  |  |  |  |  |  |  |  |  |  |  |  |  |  |
| HCW |  |  |  |  |  |  |  |  |  |  |  |  |  |  |  |
| CHW | 2.742 | 1.76 | 4.28 | 0.32 | 0.16 | 0.61* | 3.19 | 2.06 | 4.96 | 0.35 | 0.19 | 0.67 | 3.23 | 2.08 | 4.99 |
| Close relation | 1.139 | 0.78 | 1.68 | 0.83 | 0.49 | 1.41 | 1.15 | 0.79 | 1.69 | 0.90 | 0.53 | 1.52 | 1.18 | 0.8` | 1.72 |
| **Occupation** |  |  |  |  |  |  |  |  |  |  |  |  |  |  |  |
| Employed non-mine worker |  |  |  |  |  |  |  |  |  |  |  |  |  |  |  |
| Mine Worker | 0.85 | 0.6 | 1.21 | 1.75 | 1.09 | 2.82* | 0.84 | 0.59 | 1.19 | 1.55 | 0.96 | 2.48 | 0.97 | 0.68 | 1.37 |
| Unemployed | 0.74 | 0.45 | 1.22 | 1.19 | 0.59 | 2.40 | 0.73 | 0.44 | 1.2 | 1.3 | 0.66 | 2.57 | 1.07 | 0.70 | 1.61 |
| **Employment status** |  |  |  |  |  |  |  |  |  |  |  |  |  |  |  |
| Employed |  |  |  |  |  |  |  |  |  |  |  |  |  |  |  |
| Unemployed | 1.37 | 0.84 | 2.28 | 0.82 | 0.40 | 1.67 | 1.41 | 0.86 | 2.34 | 0.71 | 0.35 | 1.41 | 1.29 | 0.83 | 2.00 |
| **Phase of Treatment** |  |  |  |  |  |  |  |  |  |  |  |  |  |  |  |
| Phase 1 |  |  |  |  |  |  |  |  |  |  |  |  |  |  |  |
| Phase 2 | 0.04 | 0.02 | 0.06 | 67.6 | 36.54 | 132.64* | 0.03 | 0.02 | 0.06 | 38.32 | 23.35 | 65.44 | 0.04 | 0.02 | 0.06 |
| **TB category** |  |  |  |  |  |  |  |  |  |  |  |  |  |  |  |
| Extra Pulmonary TB |  |  |  |  |  |  |  |  |  |  |  |  |  |  |  |
| Pulmonary TB | 0.27 | 0.21 | 0.36 | 17.01 | 10.17 | 29.31* | 0.28 | 0.21 | 0.38 | 13.91 | 8.53 | 23.47 | 0.33 | 0.25 | 0.43 |
| Theta ($\theta$) | | | | 2.24 | 2.07 | 11.77 | | | | | | | | | |

**Table 6:** Random effect using Gamma frailty distribution.

|  | Weibull Gamma |  |  | Exponential Gamma |  |  | Log logistic Gamma |  |  | Lognormal Gamma |  |  | Gompertz frailty |  |  |
| --- | --- | --- | --- | --- | --- | --- | --- | --- | --- | --- | --- | --- | --- | --- | --- |
| Covariates | HR | 2.5% | 97.5% | HR | 2.5% | 97.5% | PHR | 2.5% | 97.5% | HR | 2.5% | 97.5% | HR | 2.50% | 97.50% |
| **Sex** |  |  |  |  |  |  |  |  |  |  |  |  |  |  |  |
| Female |  |  |  |  |  |  |  |  |  |  |  |  |  |  |  |
| Male | 1.12 | 0.86 | 1.47 | 1.53 | 1.29 | 1.82* | 0.76 | 0.52 | 1.12 | 0.8 | 0.54 | 1.17 | 1.13 | 0.86 | 1.49 |
| **Age category** |  |  |  |  |  |  |  |  |  |  |  |  |  |  |  |
| <20Yrs |  |  |  |  |  |  |  |  |  |  |  |  |  |  |  |
| 20-59 Yrs. | 0.71 | 0.44 | 1.18 | 114.03 | 109.62 | 120.8 | 1.17 | 0.59 | 2.29 | 1.32 | 0.68 | 2.55 | 0.75 | 0.45 | 1.22 |
| >=60 Yrs. | 7.7 | 4.72 | 12.94* | - | - | - | 0.02 | 0.01 | 0.04* | 0.02 | 0.01 | 0.05* | 8.41 | 5.1 | 13.86 |
| **Treatment contact** |  |  |  |  |  |  |  |  |  |  |  |  |  |  |  |
| HCW |  |  |  |  |  |  |  |  |  |  |  |  |  |  |  |
| CHW | 2.71 | 1.71 | 4.31* | 1.03 | 0.77 | 1.38 | 0.36 | 0.19 | 0.71* | 0.4 | 0.21 | 0.78* | 3.21 | 2.03 | 5.07 |
| Close relation | 1.12 | 0.76 | 1.68 | 0.78 | 0.64 | 0.98* | 0.93 | 0.54 | 1.61 | 1.01 | 0.58 | 1.75 | 1.15 | 0.77 | 1.71 |
| **Occupation** |  |  |  |  |  |  |  |  |  |  |  |  |  |  |  |
| Employed non-mine worker |  |  |  |  |  |  |  |  |  |  |  |  |  |  |  |
| Mine Worker | 0.98 | 0.69 | 1.39 | 0.23 | 0.21 | 0.24* | 1.70 | 1.05 | 2.74* | 1.5 | 0.93 | 2.43 | 0.98 | 0.69 | 1.4 |
| Unemployed | 1.09 | 0.72 | 1.66 | 2.06 | 1.65 | 2.48* | 0.70 | 0.38 | 1.31 | 0.87 | 0.47 | 1.59 | 1.09 | 0.72 | 1.65 |
| **Employment Status** |  |  |  |  |  |  |  |  |  |  |  |  |  |  |  |
| Employed |  |  |  |  |  |  |  |  |  |  |  |  |  |  |  |
| Unemployed | 1.21 | 0.79 | 1.89 | 0.5 | 0.45 | 0.58* | 1.25 | 0.64 | 2.45 | 0.95 | 0.49 | 1.86 | 1.27 | 0.81 | 1.97 |
| **Phase of treatment** |  |  |  |  |  |  |  |  |  |  |  |  |  |  |  |
| Phase 1 |  |  |  |  |  |  |  |  |  |  |  |  |  |  |  |
| Phase 2 | 0.04 | 0.02 | 0.07* | 11.61 | 10.69 | 12.28* | 71.16 | 37.4 | 142.96* | 39.53 | 23.61 | 68.84* | 0.04 | 0.02 | 0.06 |
| **T.B Category** |  |  |  |  |  |  |  |  |  |  |  |  |  |  |  |
| Extra Pulmonary TB |  |  |  |  |  |  |  |  |  |  |  |  |  |  |  |
| Pulmonary TB | 0.31 | 0.24 | 0.41* | - | - | - | 16.76 | 10 | 28.81* | 13.87 | 8.48 | 23.42* | 0.33 | 0.25 | 0.43 |
| Theta ($\theta$) | | | | | | | | <0.001 | | | | | | | |

Estimation of the Gamma parameter of the log logistic distribution was negligible. The variance explaining the unobserved effect is represented by the theta (*θ*) parameter. The theta (*θ*) is the shared parameter of the TB patient indicating that those grouped into the same catchment area may have similar characteristics or share the same frailty but differ between or from catchment area to catchment area. In other words, the probability of TB mortality may be similar within a cluster but different between clusters due to some characteristics that were not or could not be measured. We observed a catchment specific and overall, statistically significant difference of the unobserved effect, implying significant difference for probability among TB patients from cluster to cluster.

The estimated variance associated with the frailty effect in the log logistic regression with Gaussian frailty model presented in Table 5 was 2.24 which can be as low as 2.12 and as high as 11.13. The variance structure was found to be statistically significant indicating that survival risks among TB patients vary because of unobserved effects which is shared by patients from the same catchment regions. The estimated frailty as shown in Table 5 with Gaussian specifications indicates that a TB patient’s death in a particular catchment region increases the risk of the death for the index TB patient by 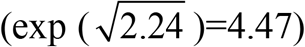 times relative to the overall risk of TB patient’s death.

### Determining Heterogeneity across Catchment Areas

However, the evidence presented in Table 6, which included the Gamma frailty parameter, shows that the frailty variance was less than 0.001. This variance value was negligible even though it was found to be statistically significant. The results show similar hazard ratios for all the covariates evaluated with confidence intervals that are close for both frailty log logistic models with Gaussian and Gamma model. This shows that there is less impact of the frailty term on parameter estimates and does not change the significance of any of the parameter estimates in the model except for those who work as miners whose hazard ratio reduced from 1.73, Cl:1.07-2.78 to 1.70, CI:1.05-2.74 when the frailty component was accounted for. However, when the frailty distribution was changed from Gamma to Gaussian the magnitude of the effect increased to HR: 1.75 (Cl: 1.09-2.82). Similarly, the magnitude of the effect of the determinants included as covariates in the model is unchanged in the presence of the random term known as frailty. The credible interval from Gamma frailty model were consistently narrower compared to the one from Gaussian frailty modelling framework in a marginally manner.

The effect of frailty models can be demonstrated by looking at the effect of exponential regression estimates for sex, occupation, and employment status on the risk of death of the T.B patient. Results from the standard model for these covariates showed no significant effect. However, the effects changed after including the Gamma frailty term in the modelling framework. Conversely, though the two modelling strategies produced comparable results, conflicting results were also observed. For instance, changing the baseline likelihood from log logistic to exponential changed the significance of covariates. Inclusion of Gamma term revealed significant increased risk of dying (HR:1.53, Cl:1.29-1.82) compared to the standard model which revealed sex not to be a significant determinant of TB mortality.

Similarly, working as a miner which was found not to be significant determinant of TB death in the standard model, became significant after adjusting for the frailty parameter (HR:0.23, Cl:0.21-0.24). Having a reduced effect, was also a surprising result as the estimates from the log logistic regression showed the opposite as shown in Tables 4, 5 and 6.

The estimates for treatment category and drug resistant exhibited consistently ambiguous risk values for all the models. The two variables were dropped and only reported in descriptive table. There were also considerable changes in the effects of selected covariates used in the Gaussian model when analyzed using Gamma frailty approach. For example, the effect of working as a miner on the risk of death became apparent as the credible interval became wider in the Gamma frailty model compared to what was reported in the Gaussian frailty model.

## Discussion and conclusion

The central question of this study was to quantify the distribution of the TB data from Lesotho and identify risk factors associated with TB mortality, which go beyond individual factors, and extend to include other factors such as the catchment areas. These factors were assumed to be best captured by assuming frailty-varying processes. In doing so, we applied a novel Bayesian framework which permitted estimation of risk at individual, and catchment area (facility) level in a unified Bayesian framework.

Our modelling approach can be considered as an extension of the traditional survival model and can be classified as a mixed effect model. These types of models have a complex structure, which is easily exploited using the Bayesian approach. Despite the complexity of our approach the results obtained from our approach are consistent with what has been reported previously (9-12).

The Kaplan–Meier survival curves show that the time to death of TB patients was different among age category, treatment contact group, employment status, phase of treatment, type of TB, and catchment areas (facility). However, clear differences were not observed among covariate such as gender, and treatment category. This result is consistent with another study which reported that age, HIV, chronic kidney disease, stroke, cancer, and chronic liver disease and cirrhosis were significant risk factors for death (23).

The log-rank tests also showed that there was a significant difference among the employment status. Similarly, the Kaplan–Meier survivor estimates for age category, treatment contact group, employment status, phase of treatment, type of TB, and catchment areas (facility) showed significant differences. These findings are consistent with those of one study done in Ethiopia (24),which found that the survival experience of patients in different categories of tuberculosis differed significantly.

Similarly, our study showed that mortality due to TB was indirectly associated with increasing age. Our study revealed that in the older TB patients, the mortality was significantly reduced. This finding was contradicting the findings reported in Tanzania (25) which found that advanced age ≥ 45 years increased the risk TB mortality. This is expected especially where data from other countries were analyzed using semi parametric approaches without necessarily testing for the proportionality of the hazard of mortality and ascertaining the distribution of the frailty parameter from the analyzed data. Nevertheless, this result was consistent with what was previously reported (26).

Our results showed that working as a miner elevates the risk of dying with TB. Similar findings have been observed (27). In their study, it was noted that TB was a leading cause of death among mineworkers and former mineworkers, causing over 20% of deaths overall. Evidence of increased TB related deaths, exceeding the proportion of deaths due to mining accidents in the mining population, were also commonly observed. Other studies have also noted increased risk of TB mortality among miner workers. For instance, Lyatuu et al reported that relative to non-miners living in the same communities, mining workers had twice the risk of mortality associated with TB (28). Data from our study were also consistent with results reported elsewhere (29), which showed that mine activities continue to be associated with a large TB mortality burden despite major efforts to ensure the safety in mining communities. This may indicate that mining worker’s TB patients usually have lower survival probability than non-mining workers TB patients. This finding is very striking and may need to be investigated further using either prospective designs or any designs that can ensure proper attribution.

Further, according to the result obtained from the multivariable log logistic–Gamma shared frailty model, the risk of death was higher in phase 2 of treatment category of TB patients relative to phase 1 and was statistically associated with the time to death of TB patients during treatment. This finding is consistent with those of a study conducted in the southern region of Ethiopia (30).

The risk of death for extra pulmonary TB patients was significantly different from that of pulmonary TB patients. This result is consistent with that of the study that was conducted in Ethopia (30) including other studies that found type of tuberculosis (pulmonary, and extra pulmonary tuberculosis) to be significant predictive factors of mortality of tuberculosis patients (31).

Interestingly, in our study, gender, was found not to be a statistically significant factor affecting the survival of TB patients. This result is consistent with the studies that were conducted in Uganda and Brazil (32, 33) but contradicts the findings of other studies that were conducted in Finland, Brazil and Singapore (34–36) that have found male TB patients to be at higher risk of dying due to TB. The differences noted could be attributed to a number of reasons including use of fixed effect models without accounting for catchment area heterogeneity and arbitrary choice of distribution for the hazard function.

The multivariate analysis was conducted using the log logistic model with Gamma frailty approach. The log logistic Gamma frailty model was arrived at after a comparison was made with other parametric Gamma frailty models as illustrated earlier in the methods section. Discrimination and final selection of the best model (log logistic) for this dataset was made using Watanabe Akaike criterion. Using the results from both Gamma and Gaussian, it is evident that heterogeneity affected significance of the determinants for TB mortality. The results showed a statistically significant community level effect on the risk of dying and demonstrated variations from catchment area to catchment area especially for results from Gaussian distribution. This approach is similar to those carried out by other researchers previously (5, 11, 12).

## Strength and Limitations

The findings as presented show a further step toward an improved understanding of the risk of TB survival among the people of Lesotho. This study has provided further evidence for the argument that information about individual patients and region is important in understanding inequalities in TB mortalities and that the application of robust statistical methodologies is also possible. The Bayesian Gamma model, the Gaussian frailties distribution specification did not exhibit any problem with the fit of the data. This shows that though the Gaussian had higher DIC values, the approach exhibits a good fit to the data when modelled through the Weibull proportional regression with frailty.

This study had a number of limitations that potentially affected the interpretation of the findings. We feel that the strength of this study would have been improved if the number of covariates were included in the modelling framework. Lack of other important covariates such as marital status, education, catchment level factors such as cultural, perception, practice and spatial information affected the ability of the model to explain the mortality risk factors acting at higher level than household. Spatial information is needed to capture temporality which might have helped to account for possible spatial effect.

Conflicting evidence especially in terms of ambiguous estimates obtained on the risk of death revealed inherit differences that could be attributed to differences in the two distributions such as Gaussian and Gamma frailty distribution specified. The differences noted need to be investigated using other Bayesian software and modelling strategies such as Stan (Hamiltonian Monte Carlo), MCMC (Markov Chain Monte Carlo Simulation) variates such as Rjags, and R2winbugs. INLA being a deterministic strategy, though fast, required different parametrization of the likelihood and the prior distribution to specify the Gamma frailty which is a non-Gaussian distributed parameter. We feel that it is this parametrization that could have affected the results especially in instances where ambiguous values were obtained. Due to the large values obtained for the DIC via the Bayesian fitted models, we anticipate model fitting issues, so our estimates should be interpreted with caution. Furthermore, ambiguous, and unrealistic estimates obtained in the Bayesian approach revealed lack of data support to the proposed Gamma frailty distributed model even though, the DIC value was small.

## Data Availability

All data produced in the present work are contained in the manuscript

## Author Contributions

IF, PSN contributed to the study concept, designed, performed the statistical analyses, and drafted the manuscript; VN, MR, LM, SM and MM participated in the data collection and data quality control; VN, MR, PSN participated in interpretation of study results; PSN, VN, MR, MO, SM, MM, RR and JD reviewed the drafts of the manuscript for critical intellectual content. All authors approved the final manuscript.

## Data Availability Statement

The datasets analyzed in this study are available from the corresponding author on reasonable request.

## Ethics Approval

The study was approved by the Lesotho Ministry of Health Research Ethics Committee and the Human Research Ethics Committee of Stellenbosch University.

